# Integrating Protein-protein Interaction Networks and Machine Learning to Identify Biomarkers of Cancer Onset

**DOI:** 10.1101/2025.11.21.25340742

**Authors:** Hongyue Chen, Min Ma, Haoyu Liu, Qian Yang, Jiqiu Wang, Jie Zheng

## Abstract

Recent large-scale plasma proteomic studies have identified a set of biomarkers for the diagnosis of early cancer onset, but the predictive performance is still a challenging problem. Most existing studies have treated proteins as independent markers, ignoring their functional interdependencies within the biological network. We consider that protein–protein interaction (PPI) networks can capture coordinated biological signals to enhance the predictive performance. We identified 1,605 high-confidence PPI pairs of proteins (corresponding to 1,155 unique proteins) from the STRING database (confidence scores>0.9). The plasma proteomic data of these pairs were extracted from a subset of 38,585 UK Biobank participants with Olink measurements (noted as UKB-PPP). The univariate Cox regression (p<0.05) integrated with elastic-net machine learning models was used to build PPI predictive model on 23 cancer types, which seven cancer types with robust PPI associated with were included in the final predictive model. In general, models included proteomics features outperformed those based on age, sex, and lifestyles. Incorporating PPI-derived interaction features further improved prediction performance in three of the seven cancer models, with melanoma showing a significant improvement compared to the base model in C-index (ΔC-index = 0.13). In summary, integrating PPI networks with proteomic models could provide predictive gains in specific cancer types and underscore the value of molecular interaction patterns as complementary biomarkers for cancer onset.

## Introduction

Cancer is one of the leading causes of death world-widely. Nearly 20 million new cancer cases were reported in 2022, along with approximately 9.7 million cancer-induced deaths (1). As such, early cancer detection is crucial for improving patients’ survival rates with more timely and effective treatment options, reducing healthcare costs, and enhancing patients’ quality of life (1, 2). Despite the potential biomarkers for early cancer detection, most existing studies focus solely on individual proteins (3). Only one-third of biomarkers are approved for clinical use, and most protein candidates could introduce noise to discovery, limiting their clinical translation (4). In addition, differences in sample handling, assay platforms, and statistical approaches further complicate validation, making it difficult to convert proteomic predictions into reliable biomarkers (5).

Such approaches neglect the rich functions rewired by protein–protein interactions (PPIs), which reflect coordinated molecular networks and pathways (6). Incorporating network information into predictive models can enhance sensitivity and specificity towards performance, but PPI networks have rarely been systematically linked with survival-based cancer risk prediction. According to previous studies, PPI networks can provide a systematic overview of cellular function by mapping how proteins physically or functionally associate. Such network-level representations can reflect biological processes that are not apparent when analyzing proteins individually (7, 8). In studies investigating cancers, oncogenic proteins and tumor suppressors often occupy vital roles in the network, suggesting that their existence can rewire oncogenic pathways. Computational and graph-theoretic analyses have further shown that oncogenic genes often correspond to key network nodes with distinct topological roles, and that modules of interacting proteins can outperform single protein molecules in diagnostic or prognostic tasks (9, 10).

However, prediction models have not yet considered PPI information, which could help enhance the biological pathway, capture additional changes in the human body compared to single protein biomarkers (11, 12). To fill in this research gap, our study aims to integrate PPI-derived features into predictive models, which may capture underlying biological characteristics and be more effective than applying protein features solely. Briefly, we applied data from the UK Biobank Pharma Proteomics Project (UKB-PPP) -the largest proteomic experiment to date, together with high-confidence, and scored PPIs from the STRING database (13) in a survival analysis for primary protein pair selection. Then, we developed a series of survival models: (1) basic models including age and sex, (2) lifestyle models further incorporating alcohol consumption, body mass index (BMI), smoking status, physical activity, and town-send deprivation index (TDI), (3) protein models adding individual proteins on top of the lifestyle models, and (4) interaction models further incorporating significant PPI pairs identified via univariate Cox regression selection. With an elastic-net Cox model and 10-fold cross-validation, we assessed whether interaction-level features can provide a predictive insight for early cancer detection across various cancer types.

## Results

### Study Cohort and Demographic Characteristics

UKB-PPP included 54,219 participants. We applied in-house quality control by excluding participants based on missing covariates in prediction models and proteomics data, and those diagnosed with cancer prior to UK Biobank baseline. In this study, we analyzed the remaining 38,585 participants aged 37 to 71 years old. 23 cancer types with at least 100 cases were included as outcomes for this study, following the workflow summarized in ***Figure 1***. The overall mean (SD) age was 57.5 (8.1) years, and 53.72 % of the participants were female.

**Figure 1.**
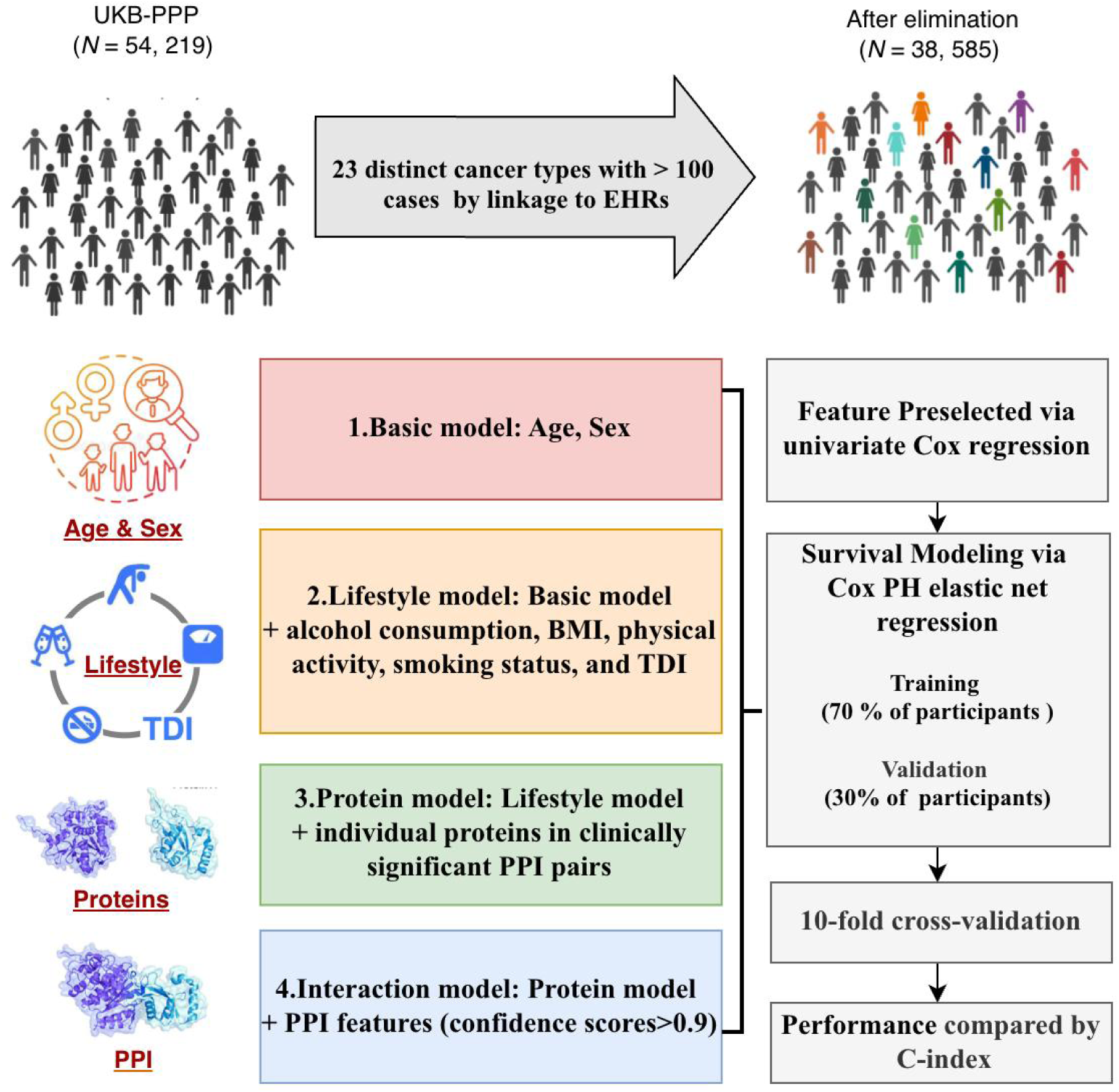
Study design to construct machine learning prediction models to identify biomarkers of cancer onsets. Participants from the UKB-PPP cohort were filtered to identify 23 cancer types with >100 cases, resulting in 38,585 individuals for analysis. Four hierarchical models were constructed: (1) Basic (age, sex); (2) Lifestyle (Basic + lifestyle); (3) Protein (Lifestyle + individual proteins from significant PPI pairs); and (4) Interaction (Protein + high-confidence PPI features). Features were prescreened through univariate Cox regression and Cox PH elastic net regression with 70/30 train–validation split and 10-fold cross-validation. Predictive performance was evaluated using the concordance index (C-index).

Among our participants, 23.4% of them had a BMI greater than 30kg/m^2^. Over half of the participants (53.33%) reported being never-smoker, while 36.72% were former smokers, and 10.40% were current smokers. Alcohol consumption patterns showed that the vast majority (93.15%) reported never drinking, while 3.68% were former drinkers and 3.17% were current drinkers. The median TDI was -2.23 (IQR: -3.69 to 0.36), with scores ranging from -6.26 to 10.00. The median physical activity was 3 times per week (IQR: 2 to 5).

### Overview of the Included Cancer Types

Referred to the International Classification of Diseases information, we initially identified 7,240 incident cancer diagnoses from 24 cancer types after UK Biobank baseline. Since our study focused on specific cancer types, we excluded the Cancer (General) category to avoid overlap and ensure each case was attributed to a defined malignancy. Therefore, 23 distinct cancer types, each of which included over 100 cases (***Figure 2***), were considered for individual PPI feature selection along with survival analyses. Among these cancer types, the most frequent diagnosis was melanoma (2,996 cases), followed by basal cell carcinoma (2,672), prostate cancer (1,213), breast cancer (940), and colorectal cancer (708). Several additional cancers, including pancreatic, kidney, esophageal, uterine, and others, were included descriptively but excluded from the prediction model due to the limited number of cases.

**Figure 2.**
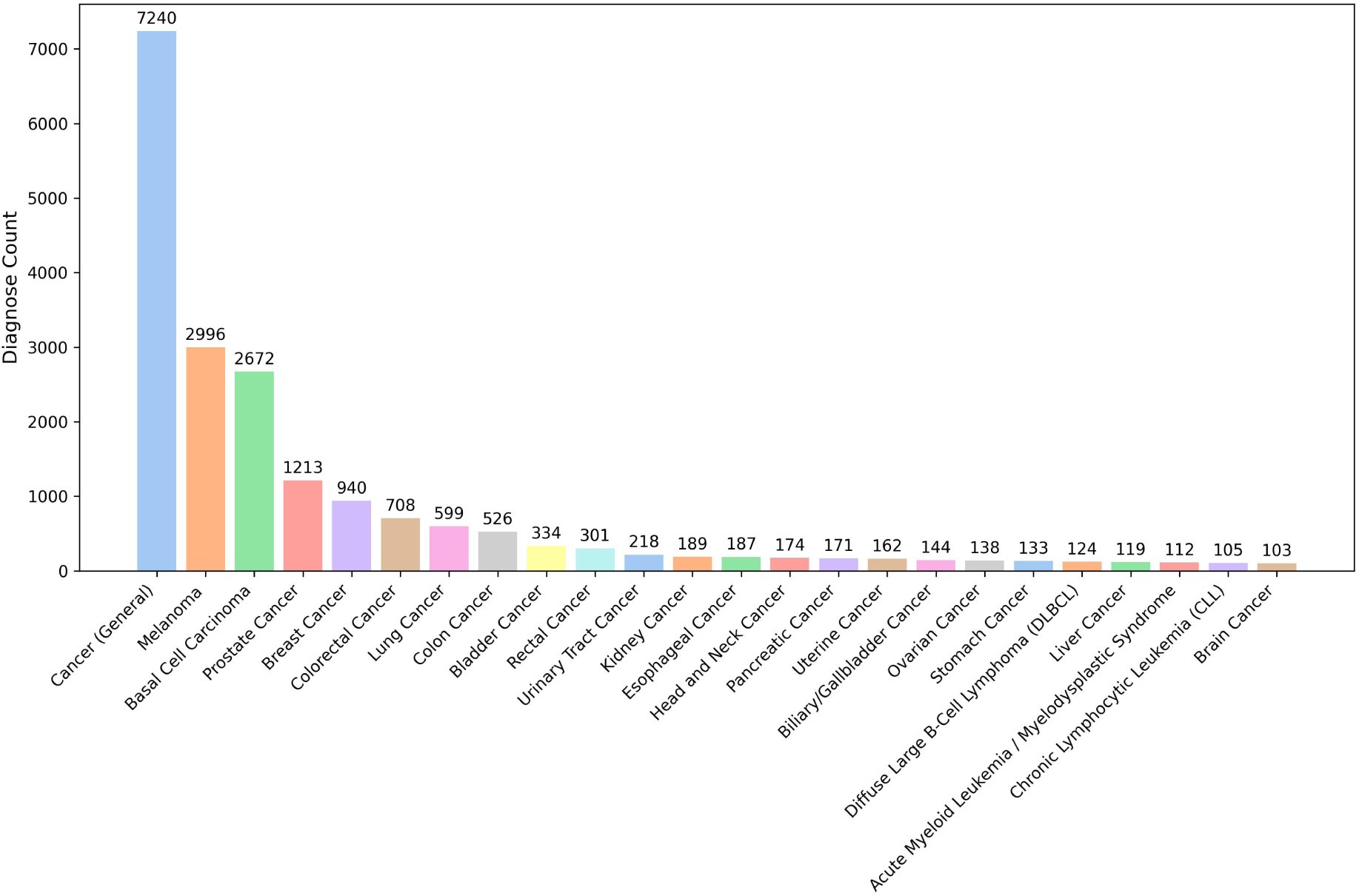
Distribution of major cancer types identified in this study.

### Summary of the Selected Protein-Protein Interactions

To gain biological insights from proteins, 2920 proteins that passed in-house quality control were searched in the STRING database to identify all available PPI networks, and we kept only high-confidence interaction pairs with a confidence score >0.9. This resulted in 1,605 PPI pairs, corresponding to 1,155 unique proteins, that could be included for the primary selection. We selected clinically significant PPIs based on the performance of individual Cox regression survival analyses (p of the interaction term <0.05). These PPI-derived predictors were evaluated alongside baseline age, sex, lifestyle covariates, and individual protein measurements.

In the pan-cancer analysis, we identified 224 significant PPI pairs associated with overall cancer risk after univariate Cox screening. As shown in ***Figure 3***, the volcano plot highlighted the distribution of these interaction features by their effect size and statistical significance.

**Figure 3.**
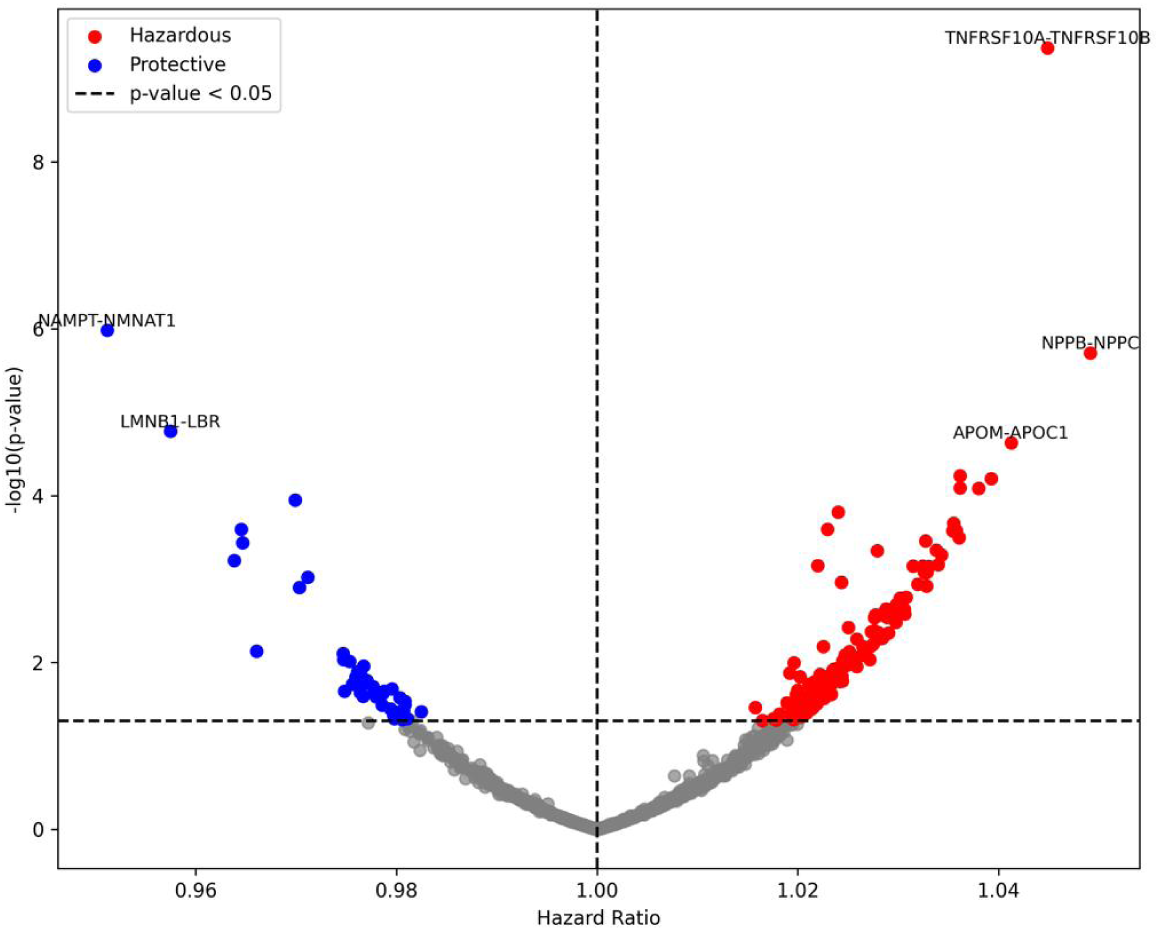
Volcano plot of significant protein–protein interactions (PPIs) associated with cancer risk. Each point represents a PPI feature, plotted by hazard ratio (x-axis) and –log10(p-value) (y-axis). Red points indicate hazardous interactions (Hazard Ratio (HR) > 1), and blue points indicate protective interactions (HR < 1). Grey dashed lines mark the significance threshold (p < 0.05) and HR = 1.

Across the 23 cancer types examined, nine showed significant PPI pairs associated with clinical outcomes (p < 0.05) in univariate Cox regression analysis. Among these, seven cancers (i.e. basal cell carcinoma, breast cancer, diffuse large B-cell lymphoma (DLBCL), liver cancer, lung cancer, melanoma, and prostate cancer) had enough case counts (>110 cases) to support stable downstream feature selection using Cox PH elastic net regression. The high-confidence PPI features retained after this selection process were depicted in ***Figure 4***, which illustrated the significant interaction networks incorporated into the final interaction model. In this network visualization, each node represented an individual protein, and each edge denotes a significant interaction supported by prior network evidence, with colors indicating the cancer types in which the PPIs were selected.

**Figure 4.**
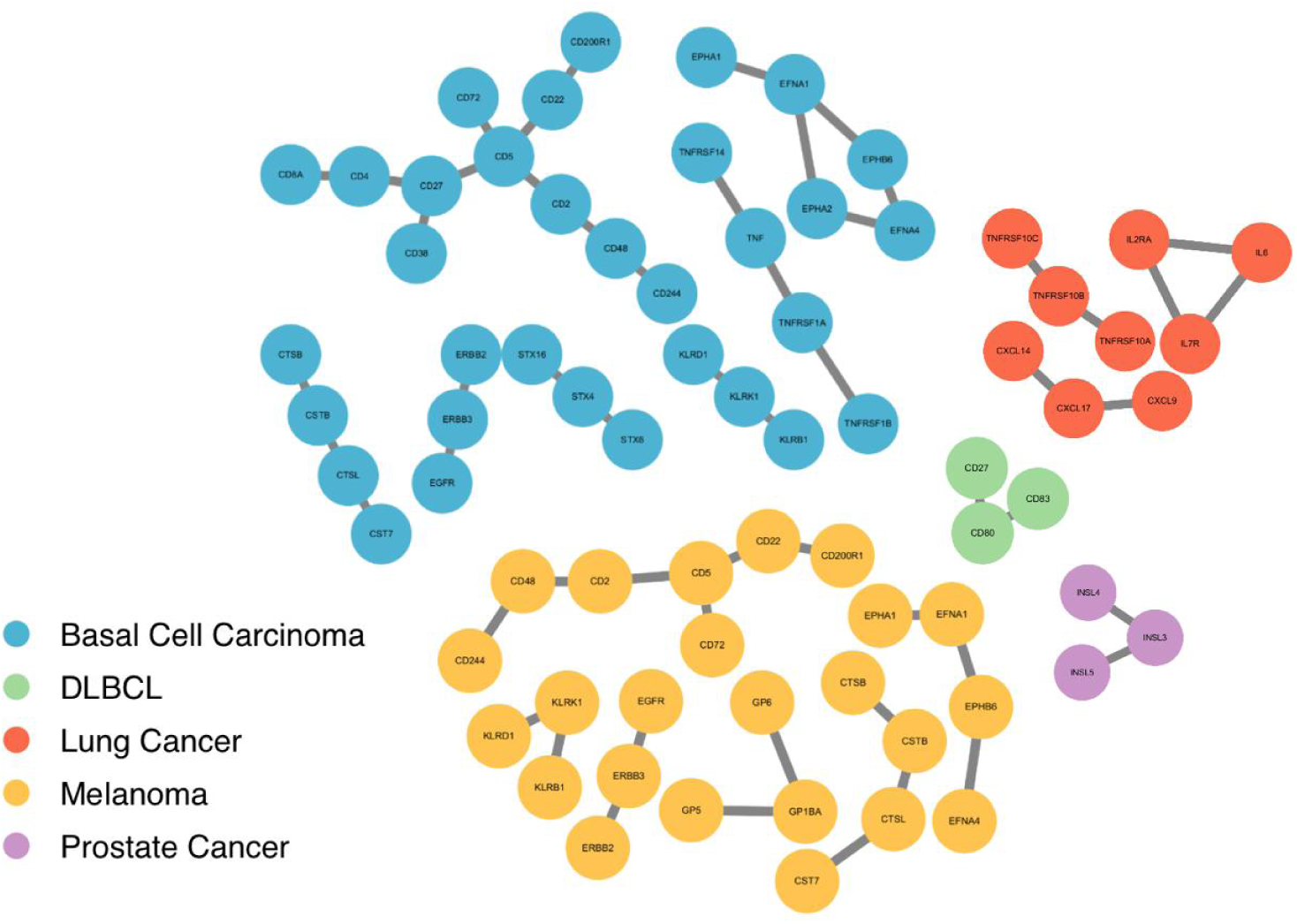
Significant protein–protein interaction (PPI) networks incorporated into the cancer risk prediction models. Cytoscape visualization of the selected high-confidence PPIs retained after feature selection. Each node represents an individual protein, and each edge denotes a significant interaction supported by network evidence. Colors correspond to cancer types for which the PPI features were selected: Basal Cell Carcinoma (blue), DLBCL (green), Lung Cancer (red), Melanoma (yellow), and Prostate Cancer (purple)

### Performance of Hierarchical Machine Learning Prediction Models

For the seven cancers with significant PPI features, we developed four Cox proportional hazards models to evaluate the incremental predictive value of proteomic and network-based features for cancer onset. First, our basic model included only age and sex, representing standard demographic risk factors. Second, our lifestyle model extended the basic model by incorporating lifestyle covariates known to influence cancer risk, including BMI, smoking status, alcohol consumption, physical activity, and the TDI. Third, our protein model further added individual protein abundances measured in plasma via the Olink platform, allowing assessment of single-protein contributions beyond demographic and lifestyle factors. Finally, our PPI model extended the protein model by including significant PPI pairs identified in univariate Cox regression (p < 0.05), representing functional interactions between proteins that may capture coordinated biological processes contributing to cancer onset.

All models were trained in 70% of the participants and evaluated in the remaining 30% of the participants. The 10-fold cross-validation was conducted to minimize model overfitting, with the participants being randomly split into ten groups. The predictive performance was quantified using the concordance index (C-index), which reflects the ability of the model to correctly rank participants by their risk of incident cancer. This approach allowed us to directly compare the incremental benefit of including lifestyle, protein, and PPI features on top of the basic prediction model across multiple cancer types.

According to model performance, the inclusion of protein features substantially improved model performance compared with basic and lifestyle models (***Figure 5***). Further extension of significant PPI pairs led to additional gains for three of the seven cancers (***Table 1***).

**Figure 5.**
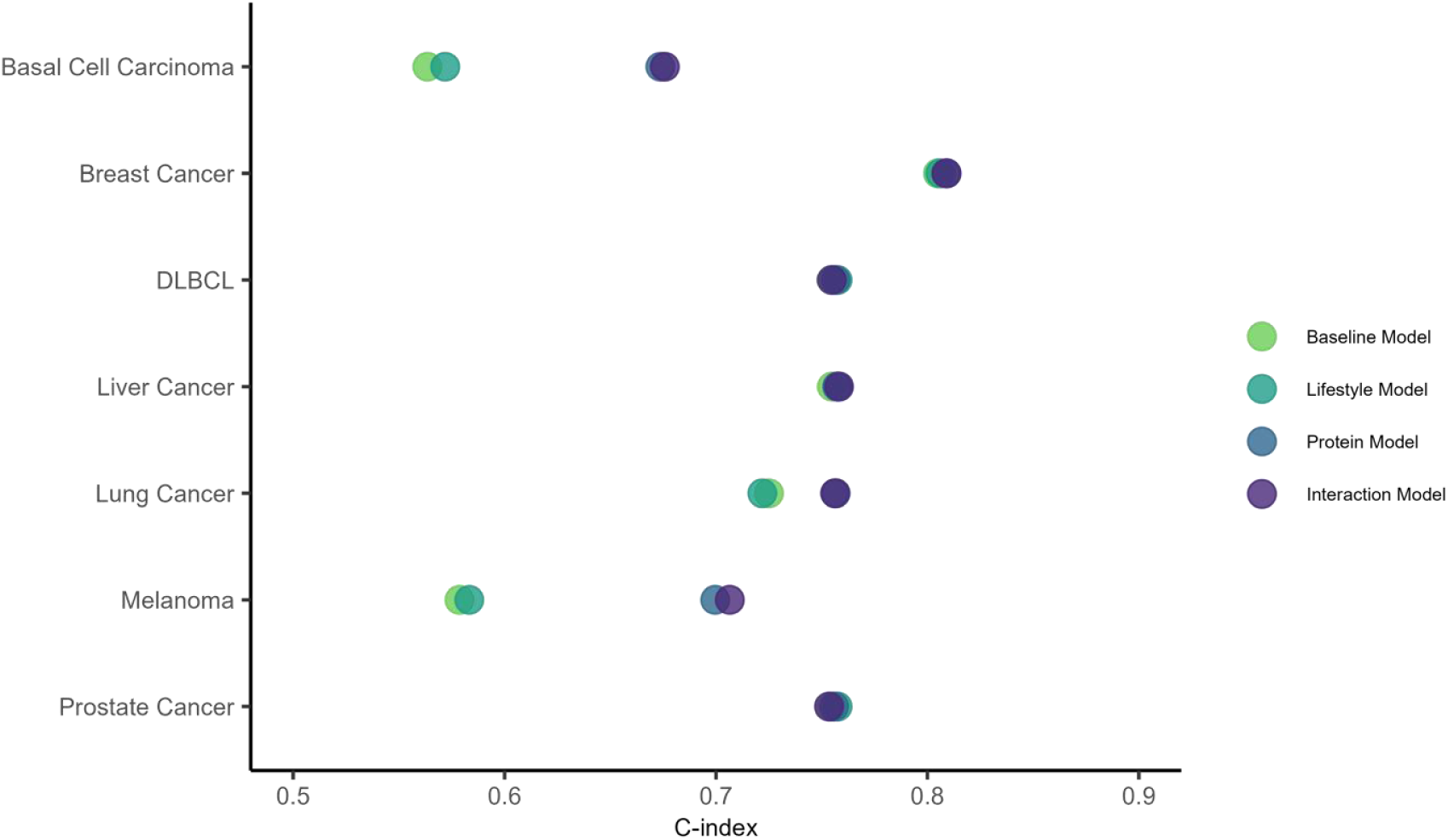
Model performance across cancer types: concordance index for nested survival models.

**Table 1.** Model performance across cancer types: concordance index for nested survival models using 10-fold cross-validation.

|  |  |  |  |  |
| --- | --- | --- | --- | --- |
| Basal Cell Carcinoma | 0.563500 | 0.571900 | 0.673700 | 0.675800 |
| Breast Cancer | 0.805000 | 0.806100 | 0.809000 | 0.809000 |
| DLBCL | 0.754700 | 0.757500 | 0.756500 | 0.754800 |
| Liver Cancer | 0.754800 | 0.757600 | 0.757300 | 0.758200 |
| Lung Cancer | 0.725000 | 0.721900 | 0.756600 | 0.756300 |
| Melanoma | 0.578700 | 0.583300 | 0.699600 | 0.706500 |
| Prostate Cancer | 0.754700 | 0.757500 | 0.755900 | 0.753500 |

We compared the predictive performance of the four models with time-dependent receiver operating characteristic (ROC) curves. Using Melanoma as an example, the area under the curve (AUC) progressively increased with the addition of lifestyle factors, individual proteins, and PPI network features in ***Figure 6***. The basic model and lifestyle model showed considerably lower discrimination (AUC = 0.58 in the validation dataset). Adding individual protein features further increased the AUC (AUC = 0.70), and the full interaction model, which integrated high-confidence PPI networks, achieved the highest predictive accuracy (AUC = 0.71). These results indicated that stepwise incorporation of biologically informed features enhances the model’s ability to distinguish individuals at a high risk of melanoma.

**Figure 6.**
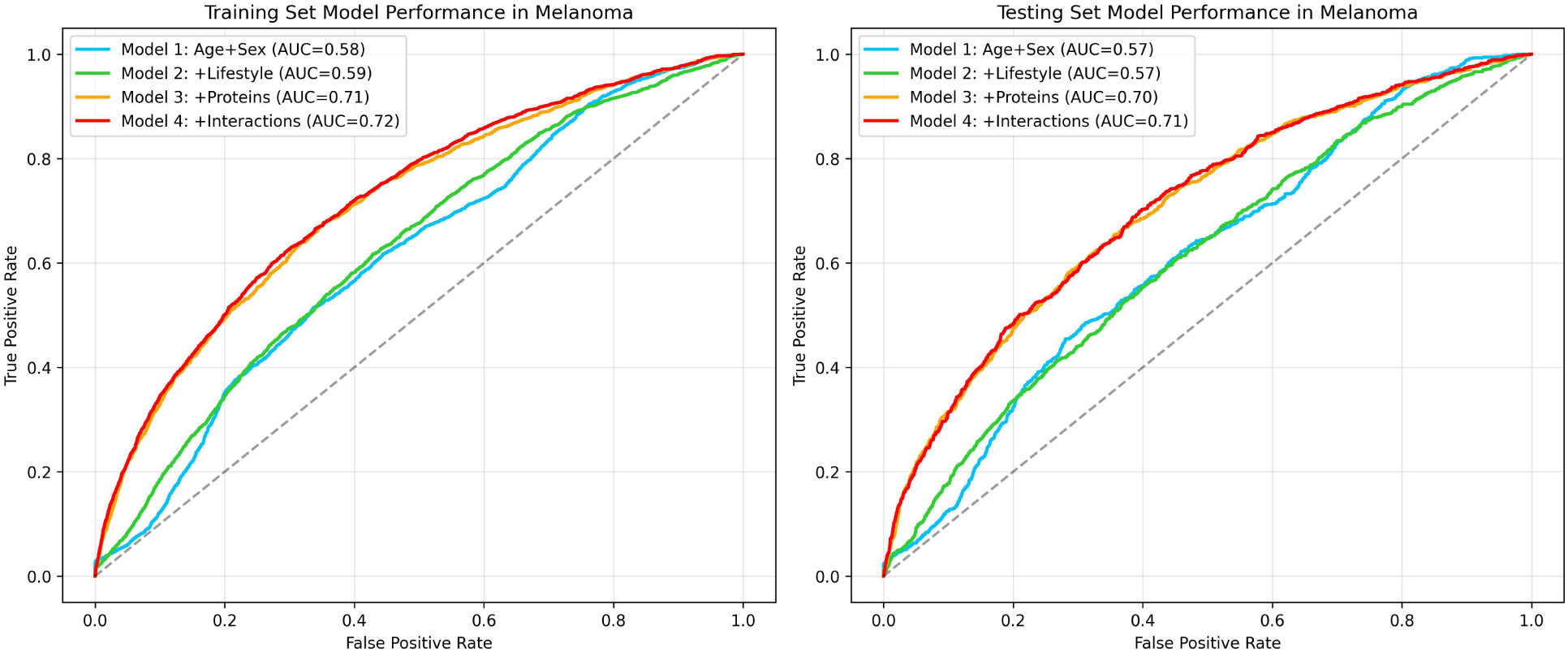
Area Under the ROC Curve (AUC) for Training and Validation Cohort.

The formal NRI test showed that the PPI model outperformed the protein model for basal cell carcinoma and melanoma (0.67 to 0.68 for basal cell carcinoma and 0.70 to 0.71 for melanoma, ***Table 1)***, indicating that PPI features captured complementary risk information not reflected by individual proteins. Liver cancer also showed a mild improvement with the interaction model (***Table 1***). For the remaining four cancers, interactions had minimal impact on predictive performance in some cases, slightly reduced the C-index compared with the protein model.

### Selected PPI Features Identified by the Interaction Model

Feature selection using Cox PH elastic-net regression also identified important PPI pairs contributing to oncologic progression. For basal cell carcinoma, top interactions included immune-related and extracellular matrix proteins, such as CD38-CD27, KLRB1-KLRK1, and COL6A3-COL3A1. In melanoma, notable interactions included CD38-CD27, ERBB3-EGFR, and NCR1-CR3LG1. For Liver cancer, selected interaction pairs were NFASC-NRCAM and COL5A1-COL4A1. These selected interactions highlighted the functional relationships that might influence cancer onset, and suggested that network-based features could be complementary to individual protein measurements in risk prediction models.

## Discussion

Across the seven cancer types analyzed in this study, the predictive accuracy steadily increased as more biological information (i.e. proteins and PPIs) was included in the models. The basic models that included only age and sex showed the lowest C-indices, implying limited predictive accuracy of applying demographic factors alone. Adding lifestyle covariates provided slight improvements compared to the basic model, which was consistent with the known contributions of behavioral and socioeconomic factors to cancer risk (14, 15). Our study showed that PPI-informed features could capture early subtle biological signals that were not evident from single protein biomarkers. This PPI-informed model performed particularly well in skin cancers, including melanoma and basal cell carcinoma, which could give us a future direction of focusing on more on skin-related cancers.

According to the results from four different models, the largest improvements were observed with integrating plasma proteomic features, which significantly improved model performance in all selected cancer types. This observation is consistent with previous prediction models using plasma proteins (16). This highlighted the value of circulating proteins as early indicators of disease-related physiological changes. For some types of cancers (basal cell carcinoma, DLBCL, liver cancers, lung cancers, melanoma, and prostate cancers), the C-indices approached or exceeded 0.70 in the protein models.

The inclusion of PPI features on top of the protein model produced varying effects across different cancer types. For basal cell carcinoma, melanoma, and liver cancer, the interaction model slightly increased the C-index compared with the protein model. The improvements suggested that certain cancer types could be influenced by coordinated dysregulation of interacting proteins rather than isolated protein level changes. These cancers might involve early or subtle molecular network perturbations that were captured by pairwise protein-protein interactions.

Elastic-net feature selection could identify some biologically plausible protein-protein interactions in the prediction model. Here are three examples in which the interaction models outperformed the protein model. In terms of basal cell carcinoma, selected interactions included immune activation pairs (e.g., CD38 × CD27, KLRB1 × KLRK1) and extracellular matrix remodeling proteins (COL6A3 × COL3A1). These were consistent with known roles of immune surveillance and stromal remodeling in skin cancer initiation (17, 18). In terms of melanoma: The presence of ERBB3 × EGFR and killer-cell ligand interactions suggests early involvement of growth factor signaling and innate immune pathways—both well-established in melanomagenesis (19). In terms of liver cancer, key pairs such as NFASC × NRCAM and COL5A1 × COL4A1 point to cell adhesion and matrix interactions, aligning with the fibrosis–inflammation axis central to liver cancer development. In summary, our results supported the concept that integrating proteomics with network biology could enhance cancer risk prediction in early stages of cancer. Proteins often interact within molecules and complexes, and disruptions within these networks might be detected earlier than strong impacts on individual proteins. PPI features might improve specificity, reducing noise from the thousands of proteins measured. Cancer types with strong extracellular matrix or immune involvement appear particularly amenable to interaction-based modeling(20).

In breast cancer, DLBCL, lung cancer, prostate cancer, adding interaction terms did not improve performance. Several possible speculations could explain the results. First, protein-protein interactions might not be strong indicators in early disease development of those cancers. Early tumorigenesis did not always produce changes across interacting proteins. In certain cancer types, early biological alterations might occur at the level of genetics, epigenetics, or subtle signaling events that were not yet detectable in circulating proteins (21). Second, the individual protein information might already reflect the key biological changes associated with early disease. These proteins could act as strong markers on their own, capturing the primary molecular signals driving cancer onset. As a result, adding PPI features might provide little additional predictive information, because the model has already incorporated most of the relevant signal (22). Third, the insufficient cases caused elastic-net model to exclude some useful interaction features. Even though the interaction pairs actually were crucial for tumorigenesis, the sparsity constraints could still filter out these features because they didn’t have enough evidence to keep the features (23). Finally, some interactions might only reveal their predictive value when considered within broader pathways rather than as isolated pairs. Biological processes often involve coordinated activity among multiple proteins, so analyzing only pairwise edges could miss these higher-order dependencies (24).

Taking advantage of a high-quality PPI database, our prediction model incorporated unique, high-confidence PPIs, substantially increasing the number of biologically informed predictors compared with models based solely on individual proteins. By integrating PPI networks, our models not only account for the presence of individual proteins but also consider their functional interactions, capturing complex biological relationships that may better reflect the mechanisms underlying cancer risk. This network-based approach allows identification of interaction-driven biomarkers that could be overlooked by traditional single-protein models, enhancing the sensitivity and interpretability of the predictive framework.

This study still had several limitations. First, although incorporating PPI features could provide additional predictive insights for some cancers, the improvements were modest and only for special cancer types, suggesting that simple pairwise interactions may capture only a small fraction of the true biological complexity of protein networks. Second, for some cancer types in the study population, the number of incident cases was relatively small, which limited statistical power and might have prevented reliable selection of meaningful interaction features in the prediction model. Third, the current release of the Olink proteome data from UKB-PPP project measured 2,920 proteins, with 1,155 proteins overlapping with PPI features. This may partly explain why we got modest improvement in the performance of the PPI-informed model. Finally, each participant had their proteomic data measured only once on the date when they attended the study, and protein networks might rewire as early disease developed. If we had multiple proteomic measurements over several years till diagnosis, we might discover some evolving patterns that could be more informative for early cancer detection than a single snapshot. Looking ahead, our future work could be finished in four different aspects. First, manually selecting experimentally-proven interactions supported by existing studies could yield stronger and more interpretable improvements towards our prediction models, and it could reduce the sparse constraints generated by elastic-net selection. Second, extending our pairwise framework by incorporating higher-order network structures, such as triplet interactions or graph-embedding approaches that could capture broader patterns of connectivity beyond simple pairs. Pathway-informed regularization could also help feature selection in biologically coherent modules instead of isolated protein pairs. Third, integrating proteomics with other complementary data, such as methylation, transcriptomics or metabolites data, might further enhance the performance of the model. Another promising direction was the integration of interaction pairs based on biological background, rather than relying solely on statistical models. Finally, the external validation in independent cohorts would be important to establish the robustness and generalizability of these network-based biomarkers. Some of these directions are likely to be solved once the full 600K protein measurements from UKB-PPP are released.

Overall, our findings demonstrated the potential of PPI network modeling as a complementary strategy for early cancer detection. By integrating proteomic data within biological network information, we also provided a novel direction toward potentially more sensitive biomarkers. Further studies that include large-scale longitudinal proteomic measurements, integrate multiple molecular traits, and apply higher-dimensional network representations are warranted to fully evaluate and validate the utility of interaction-based biomarkers across different cancer types and populations.

## Methods

### Participants

This study leveraged data from the UKB, a nationwide cohort of approximately 500,000 adults recruited across the United Kingdom between 2006 and 2010. Among them, 462,399 European participants had linked clinical records. Our analysis utilized data from the UKB-PPP, in which proteomic profiling was performed on EDTA-plasma samples from 54,219 individuals. The UKB-PPP design comprised: (1) a random subset (n=46,595); (2) a consortium-selected group (n=6,356); and (3) a COVID-19 imaging cohort (n=1,268) (25). UKB has received ethical approval from the UK National Health Service’s National Research Ethics Service (ref 11/NW/0382), and our study was performed under the application number 104086.

Since the goal of this study was to evaluate biomarkers of cancer onset, we focused strictly on incident cancer events. Participants with any prior cancer diagnosis before the attending date were excluded to ensure that proteomic signals only reflected pre-diagnostic biology rather than consequences of known or treated disease. We further excluded individuals enrolled in the COVID-19 repeat imaging assessment, since SARS-CoV-2 infection was known to induce genomic alterations, particularly in the immune system, and cause inflammation, which could be a confounder for early cancer detection (26, 27). Patients who withdrew during the middle of the study to ensure complete follow-up information for all remaining individuals. At the same time, we also removed individuals with missing demographic or lifestyle covariates, such as age, alcohol consumption, BMI, physical activity, smoking status, and TDI. Since these lifestyle-related variables were required for our basic and lifestyle-adjusted survival models for our construction, excluding participants with incomplete information ensured that all models were trained and tested on the same set of individuals, preventing bias from inconsistent sample sizes across comparisons. After going through these procedures, the final study population included 46,304 participants of European ancestry with full demographic, lifestyle, and proteomic information.

### Proteomics Data Processing

Proteomic data were generated using the UK Biobank Olink Explore 3072 platform, across eight protein panels (cardiometabolic, cardiometabolic II, inflammation, inflammation II, neurology, neurology II, oncology, and oncology II), which initially quantified 2,923 unique plasma proteins (25). The dataset underwent multiple quality control procedures to ensure suitability for early cancer prediction, including the exclusion of outlier samples, filtering of data with quality control or assay warnings, and removal of likely sample swaps (25).

To enable reliable imputation and downstream modelling, we first applied missingness filters at both the individual and protein levels. A total of 7,719 participants with >20% missing proteomic measurements were excluded. Subsequently, proteins with >50% missingness across the remaining samples were removed, resulting in the exclusion of 3 proteins (GLIPR1, NPM1, PCOLCE) (25, 27). For the remaining data, the missing NPX values were imputed using K-nearest neighbors (KNN; k=10) imputation, and all of the values were inverse-rank normalized to reduce skewness and stabilize variance for prediction modeling (28). After quality control, 38,585 participants and 2,920 proteins remained for downstream analyses.

To incorporate biological context, these 2,920 proteins were mapped to the STRING v12.0 database to identify PPIs (13). Only the highest-confidence pairs (score >0.9) were selected, resulting in 1,605 PPI pairs and corresponding 1,155 unique proteins for primary model construction.

### Outcome Definition and Covariates

The outcome of this study was the incident of cancer. Cancer cases were identified using International Classification of Diseases codes from linked national cancer registries and hospital episode statistics. To ensure adequate statistical power and statistical robustness, we only included cancer types for which incident cases were greater than 100, so we reduced the initial 111 different cancers to 23, which excluded general cancer or repetition. Time-to-event was calculated from the baseline assessment date (the date of attending the UKB visit) to either the date of cancer diagnosis or the end of follow-up, or censoring.

Demographic information included age at the baseline assessment date and sex. Lifestyle covariates comprised of alcohol consumption, BMI, physical activity, smoking status, and the TDI. Continuous variables, such as age, BMI, and TDI were standardized by using the *StandarScaler* function from scikit-learn (v1.6.0), and categorical variables (sex, alcohol consumption, physical activity, and smoking status) were factored for Cox PH hazards models. The demographic covariates were incorporated into all models, enabling evaluation of the additional predictive value provided by proteomic and PPI features.

### PPI Feature Selection

To identify the PPI that could worsen the clinical outcomes for each cancer type, we performed univariate Cox PH regression for each interaction while adjusting for age, sex, and lifestyle covariates using *CoxPHFitter* from the *lifelines* library (v0.28.0). Interaction pairs were evaluated individually to determine their association with incident cancer.

PPI pairs with a p-value < 0.05 were considered clinically significant and marked for final prediction model construction. This approach allowed us to focus on interactions that provided independent prognostic information beyond baseline demographic and lifestyle risk factors, reducing dimensionality and enhancing model interpretability. The selected PPI pairs were then fit into Cox PH elastic net regression to assess their combined predictive contribution alongside individual proteins.

### Survival Modeling and Feature Selection

To evaluate the predictive value of proteomic and PPI features for incident cancer, we constructed four different nested Cox PH models with regularization, using *CoxnetSurvivalAnalysis* from the *scikit-survival* package (v0.23.0). The dataset was randomly divided into training and testing subsets using a 70:30 split via the train_test_split function from the *scikit-learn* library (v1.6.0). Four sets of model structures were the same for each cancer type:

1. Basic model: age and sex
2. Lifestyle model: basic model plus alcohol consumption, BMI, physical activity, smoking status, and TDI
3. Protein model: lifestyle model plus individual proteins in clinically significant PPI pairs 4.Interaction model: protein model plus selected highest-confidence PPI features, where interactions were preselected via univariate Cox regression results for each cancer type

We set the L1 ratio as 0.9 for our Cox PH elastic net regression model, so it applied a moderate amount of shrinkage to the coefficients. This approach tried to push smaller, less important features toward zero while keeping the most informative ones, which helped the model to be less overfitting and select only the features that truly contribute to predicting cancer, without being too aggressive and removing potentially useful signals (29). The penalty shrinks less informative coefficients toward zero, retaining only the most predictive proteins or interaction pairs.

### C-index calculation

The C-index was a measure of how well a survival model could correctly rank individuals by their risk, computed via *concordance_index_censored* from *scikit-survival* (v0.23.0). It reflected the probability that, for a randomly selected pair of participants, the one who experienced the event (e.g., cancer onset) earlier was also predicted to have a higher risk by the model.

We compared all possible pairs of participants where the incident time was known. A pair was concordan**t** if the participant with the shorter observed survival time also had a higher predicted risk. The C-index was then computed as the number of concordant pairs divided by the total number of comparable pairs. A C-index of 0.5 indicated no predictive ability, while a value of 1.0 represents perfect discrimination. The model with a higher C-index stated that the model had higher predictive ability.

### Cross-Validation and Model Comparison

To rigorously assess model performance and prevent overfitting, all survival models were evaluated using 10-fold cross-validation. The training set was randomly split into 10 equally sized folds. In each iteration, nine portions were used for model training and feature selection, and the remaining fold was used for validation. The procedure included repetition across all folds, and the average C-index, which was the determined factor of model performance, was calculated to summarize predictive performance. This method helped to ensure that performance estimates were robust and generalizable, minimizing bias that could arise from a single train-test split (30).

Comparisons were made across the four models, basic, lifestyle, protein, and interaction in each cancer type. This allowed us to determine the quality of a model with the addition of predictive value, such as proteomics data and PPI networks.

## Funding

This research was funded by the Noncommunicable Chronic Diseases-National Science and Technology Major Project (2024ZD0531500, 2024ZD0531502), the National Key Research and Development Program of China (2022YFC2505200, 2022YFC2505201, 2022YFC2505203), and the National Natural Science Foundation of China (32500519, 32570728). J.Z., and Q.Y. are affiliated with the Medical Research Council Integrative Epidemiology Unit at the University of Bristol which is supported by the Medical Research Council (MC_UU_00032/3, MC_UU_00032/5) and the University of Bristol. These funders had no role in study design, data collection and analysis, decision to publish or preparation of the manuscript.

## Acknowledgements

We thank the participants, contributors, and researchers of the UKB for making data available for this study. We thank the research and development teams at the 13 participating UKB-PPP companies (Alnylam Pharmaceuticals, Amgen, AstraZeneca, Biogen, Calico, Bristol-Myers Squibb, Genetech, GlaxoSmithKline (GSK), Janssen Pharmaceuticals, Novo Nordisk, Pfizer, Regeneron, and Takeda) for funding the study. All 13 companies listed as part of the UKB-PPP were involved in the generation of the proteomic data used in the present study.

## Data availability

Full information on how to access UKB data can be found at its website (https://www.ukbiobank.ac.uk/use-our-data/).

## Author contributions

H.C, M.M, and J.Z, conceptualized the study design and consulted on methods and results. H.C and J.Z carried out all analyses. H.C wrote the first draft of the paper, with all other authors interpreting the results and making important critical revisions. M.M performed quality control on the proteomics dataset and study flow chart designation. H.L cytoscape network graphic and model performance graphic. H.C, Q.Y and J.Z were consulted on methodology. All authors reviewed the approved the manuscript.

## Competing interests

All authors declare no competing interests.

## Notes

### Competing Interest Statement

The authors have declared no competing interest.

## References

1. Bray F, Laversanne M, Sung H, Ferlay J, Siegel RL, Soerjomataram I, et al. Global cancer statistics 2022: GLOBOCAN estimates of incidence and mortality worldwide for 36 cancers in 185 countries. CA Cancer J Clin. 2024;74(3):229–63.

2. Crosby D, Bhatia S, Brindle KM, Coussens LM, Dive C, Emberton M, et al. Early detection of cancer. Science. 2022;375(6586):eaay9040.

3. Barker AD, Alba MM, Mallick P, Agus DB, Lee JSH. An Inflection Point in Cancer Protein Biomarkers: What was and What’s Next. Molecular & Cellular Proteomics. 2023;22(7).

4. McDermott JE, Wang J, Mitchell H, Webb-Robertson B-J, Hafen R, Ramey J, et al. Challenges in biomarker discovery: combining expert insights with statistical analysis of complex omics data. Expert Opinion on Medical Diagnostics. 2013;7(1):37–51.

5. Frantzi M, Bhat A, Latosinska A. Clinical proteomic biomarkers: relevant issues on study design & technical considerations in biomarker development. Clinical and Translational Medicine. 2014;3(1):e7.

6. Yang L, Zhang YH, Huang F, Li Z, Huang T, Cai YD. Identification of protein-protein interaction associated functions based on gene ontology and KEGG pathway. Front Genet. 2022;13:1011659.

7. Turanalp ME, Can T. Discovering functional interaction patterns in protein-protein interaction networks. BMC Bioinformatics. 2008;9:276.

8. Cafarelli TM, Desbuleux A, Wang Y, Choi SG, De Ridder D, Vidal M. Mapping, modeling, and characterization of protein-protein interactions on a proteomic scale. Curr Opin Struct Biol. 2017;44:201–10.

9. Xiong W, Xie L, Zhou S, Liu H, Guan J. The centrality of cancer proteins in human protein-protein interaction network: a revisit. Int J Comput Biol Drug Des. 2014;7(2-3):146–56.

10. Yuan X, Chen J, Lin Y, Li Y, Xu L, Chen L, et al. Network Biomarkers Constructed from Gene Expression and Protein-Protein Interaction Data for Accurate Prediction of Leukemia. J Cancer. 2017;8(2):278–86.

11. Barabasi AL, Gulbahce N, Loscalzo J. Network medicine: a network-based approach to human disease. Nat Rev Genet. 2011;12(1):56–68.

12. Carson MB, Lu H. Network-based prediction and knowledge mining of disease genes. BMC Med Genomics. 2015;8 Suppl 2(Suppl 2):S9.

13. Szklarczyk D, Nastou K, Koutrouli M, Kirsch R, Mehryary F, Hachilif R, et al. The STRING database in 2025: protein networks with directionality of regulation. Nucleic Acids Res. 2025;53(D1):D730–D7.

14. Klein WMP, O’Connell ME, Bloch MH, Czajkowski SM, Green PA, Han PKJ, et al. Behavioral Research in Cancer Prevention and Control: Emerging Challenges and Opportunities. J Natl Cancer Inst. 2022;114(2):179–86.

15. Li S, He Y, Liu J, Chen K, Yang Y, Tao K, et al. An umbrella review of socioeconomic status and cancer. Nat Commun. 2024;15(1):9993.

16. Budnik B, Amirkhani H, Forouzanfar MH, Afshin A. Novel proteomics-based plasma test for early detection of multiple cancers in the general population. BMJ Oncology. 2024;3(1):e000073.

17. Pfisterer K, Shaw LE, Symmank D, Weninger W. The Extracellular Matrix in Skin Inflammation and Infection. Front Cell Dev Biol. 2021;9:682414.

18. Ramelyte E, Nageli MC, Hunger R, Merat R, Gaide O, Navarini AA, et al. Swiss Recommendations for Cutaneous Basal Cell Carcinoma. Dermatology. 2023;239(1):122–31.

19. Pastwinska J, Karas K, Karwaciak I, Ratajewski M. Targeting EGFR in melanoma -The sea of possibilities to overcome drug resistance. Biochim Biophys Acta Rev Cancer. 2022;1877(4):188754.

20. Yam JW, Tse EY, Ng IO. Role and significance of focal adhesion proteins in hepatocellular carcinoma. J Gastroenterol Hepatol. 2009;24(4):520–30.

21. Tlsty TD. Genetic and epigenetic changes in early carcinogenesis. Breast Cancer Research. 2005;7(2):S.14.

22. Chen H, Lai X, Zhu Y, Huang H, Zeng L, Zhang L. Quantitative proteomics identified circulating biomarkers in lung adenocarcinoma diagnosis. Clinical Proteomics. 2022;19(1):44.

23. Pavlou M, Ambler G, Seaman SR, Guttmann O, Elliott P, King M, et al. How to develop a more accurate risk prediction model when there are few events. BMJ. 2015;351:h3868.

24. Xie W, Li W, Zhang S, Wang L, Yang J, Zhao D. A novel biomarker selection method combining graph neural network and gene relationships applied to microarray data. BMC Bioinformatics. 2022;23(1):303.

25. Sun BB, Chiou J, Traylor M, Benner C, Hsu YH, Richardson TG, et al. Plasma proteomic associations with genetics and health in the UK Biobank. Nature. 2023;622(7982):329–38.

26. Zhang JY, Whalley JP, Knight JC, Wicker LS, Todd JA, Ferreira RC. SARS-CoV-2 infection induces a long-lived pro-inflammatory transcriptional profile. Genome Med. 2023;15(1):69.

27. Carrasco-Zanini J, Pietzner M, Davitte J, Surendran P, Croteau-Chonka DC, Robins C, et al. Proteomic signatures improve risk prediction for common and rare diseases. Nat Med. 2024;30(9):2489–98.

28. Gadd DA, Hillary RF, Kuncheva Z, Mangelis T, Cheng Y, Dissanayake M, et al. Blood protein assessment of leading incident diseases and mortality in the UK Biobank. Nat Aging. 2024;4(7):939–48.

29. van der Wurp H, Groll A. Introducing LASSO-type penalisation to generalised joint regression modelling for count data. AStA Advances in Statistical Analysis. 2023;107(1):127–51.

30. Malakouti SM, Menhaj MB, Suratgar AA. The usage of 10-fold cross-validation and grid search to enhance ML methods performance in solar farm power generation prediction. Cleaner Engineering and Technology. 2023;15:100664.

